# Ultra-high-resolution Photon-Counting CT for Leaflet-Specific Evaluation of Fibrosis and Calcification in Aortic Stenosis

**DOI:** 10.64898/2026.10.27.26366483

**Authors:** Mohammed Bahaaeldin, Jasmin Shamekhi, Bernardo S. Franklin, Sebastian Nowak, Alexander MC Böhner, Dmitrij Kravchenko, Jan Wrobel, Raphael Phinicarides, Lenhard Pennig, Jan P. Janssen, Carsten Gietzen, Sebastian Zimmer, Stephan Baldus, Malte Kelm, Georg Nickenig, Tobias Zeus, Victor Mauri, Julian A. Luetkens

**Author notes:** **Address for correspondence:** Prof. Julian A. Luetkens, University Hospital Bonn, Department of Diagnostic and Interventional Radiology, Venusberg-Campus 1, 53127 Bonn. **Funding:** This study was funded by the Collaborative Research Center TRR259 (German Research Foundation [DFG] project number 397484323). **Disclosures:** The authors declare no conflicts of interest.

## Abstract

**Objectives:** Ultra-high-resolution photon-counting detector (PCD)-CT may enable leaflet-specific analysis of aortic valve fibrosis and calcification, providing insights into early aortic stenosis (AS) disease progression. This study aimed to investigate how PCD-CT leaflet-specific calcific and fibrotic volumes influence cardiovascular magnetic resonance (CMR) leaflet mobility.

**Materials and methods:** Participants with moderate aortic AS prospectively underwent both PCD-CT (n=80) and CMR (n=58). Using PCD-CT, calcific and fibrotic volumes were segmented for right (RCC), left (LCC), and non-coronary cusps (NCC) and indexed to the aortic annulus area (ICV and IFV). CMR-derived leaflet mobility index (LMI) was assessed. Two raters compared image quality using a Likert-scale (1=poor to 5=excellent) and distinguishability of individual leaflets in PCD-CT with EID-CT (n=82).

**Results:** PCD-CT analysis showed that calcification was higher in males (e.g., ICV_RCC_: 15±16 vs. 4±7 µL/cm^2^;p<0.001) while fibrosis was higher in females (e.g., IFV_LCC_; 47±18 vs. 34±15 µL/cm^2^;p=0.004) across all leaflets. Fibrocalcific ratio was higher in LCC (53±276) compared to RCC (19±48;p<0.001) and NCC (13.5±34;p<0.001). LMI of RCC was higher in females compared to men (0.66±0.21 vs. 0.38±0.33;p=0.008). LMI_LCC_ and LMI_NCC_ showed no sex differences.

LMI_RCC_ negatively correlated with ICV_RCC_ (ρ=-0.64[-0.77, −0.46];p<0.001) and LMI_NCC_ with ICV_NCC_ (ρ=-0.69[-0.80, −0.53];p<0.001), whereas LMI_LCC_ negatively correlated with IFV_LCC_ (ρ=-0.35[-0.56,-0.10];p=0.007), particularly in females (ρ=-0.70[-0.89, −0.31];p = 0.003). PCD-CT yielded higher image quality (5[IQR4–5] vs. 3[3–4];p<0.001) and leaflet distinguishability (79/80[99%] vs. 42/82[51%];p<0.001) than EID-CT.

**Conclusions:** Ultra-high-resolution PCD-CT allows for leaflet-specific quantification of indexed fibrotic and calcific volume. These volumes demonstrate sex- and leaflet-specific differences that may distinctly contribute to AS pathophysiology.

**Key Points:** Detailed leaflet-level characterization of fibrotic and calcific remodeling in aortic stenosis remains limited with energy integrating CT.

Photon-counting CT enabled reliable leaflet-specific quantification of fibrosis and calcification, identified sex-related tissue distribution patterns, and demonstrated associations between calcification and impaired leaflet mobility.

Leaflet-specific photon-counting CT assessment may enhance our understanding of aortic stenosis progression and support more individualized imaging evaluation through improved differentiation of fibrotic and calcific valvular remodeling.

**Clinical Trials Registration:** NCT05851209

## Introduction

Aortic stenosis (AS) is the most prevalent valvular heart disease in the Western world and a major cause of cardiovascular morbidity and mortality[1,2]. AS is considered to be an active, complex disease process involving endothelial damage, lipid accumulation and inflammation[3]. These processes culminate in fibrosis and calcification of the valve, with distribution patterns that differ significantly between sexes[4].

Computed tomography (CT) is increasingly being utilized for AS assessment. CT calcium scoring is widely established for severity assessment[5], yet it fails to capture the non-calcific burden that contributes to AS. Recently, contrast-enhanced CT-angiography using energy-integrating detector CT (EID-CT) has emerged as a robust method for evaluating the "fibro-calcific" volume of the valve[6], offering a more comprehensive assessment of disease severity. Despite these advances, EID-CT faces technical limitations. Blooming artifacts caused by dense calcium can obscure adjacent non-calcified tissue, and limited spatial resolution often hinders analysis of fine structures.

Photon-counting CT (PCD-CT) represents a paradigm shift in cardiovascular imaging. Due to its ultra-high spatial resolution and improved spectral separation, it can significantly reduce blooming artifacts and provide superior soft-tissue contrast compared to EID-CT[7,8]. Theoretically, these unique capabilities should also enable a more detailed and advanced analysis of the aortic valve including high-quality leaflet-specific segmentations. Such precise visualization of the aortic valve could advance our understanding of AS etiology and pathophysiology. Furthermore, granular tissue characterization may be crucial step toward developing more accurate, individualized prediction models for disease progression[9]. Cardiovascular magnetic resonance (CMR) is yet another extensively evaluated imaging tool for the characterization of aortic valve morphology and function[10,11] and offers a superior temporal resolution compared with CT enabling radiation-free, dynamic assessment of valvular motion and flow throughout the cardiac cycle.

This study aimed to explore whether ultra-high resolution PCD-CT allows for granular and precise assessment of aortic valve stenosis with a leaflet-specific segmentation of fibrosis and calcification. Furthermore, we aimed to explore correlations between CMR-derived aortic leaflet mobility and indexed fibrotic and calcific volume.

## Methods

### Study design and population

This prospective study is a cross-sectional sub-analysis from the C06 project of the Collaborative Research Center TRR259 (German Research Foundation [DFG] project number 397484323; NCT05851209). C06 aims to identify and integrate morphological, functional, genetic, and immunological biomarkers to predict disease progression in participants with mild to moderate aortic stenosis. Participants were included from April 2023 to June 2026. The study was conducted in accordance with the Declaration of Helsinki and approved by the local ethics committees. All participants gave written informed consent prior to study inclusion.

Cardiac CT angiography including CT calcium scoring and CMR were performed alongside echocardiography and blood laboratory analysis. Exclusion criteria included bicuspid aortic valve anatomy, previous aortic valve replacement and endocarditis. Participants with renal insufficiency (estimated glomerular filtration rate < 30 mL/min/1.73m^2^) did not undergo cardiac CT angiography. However, participants who did not undergo CMR due to contraindications (e.g., incompatible active implants or severe claustrophobia) were still included in this study. CMR was performed at the same day as cardiac CT angiography. Participants were recruited at different centers using the same inclusion criteria, one with a PCD-CT and one with EID-CT scanner (Figure 1).

**Figure 1.**
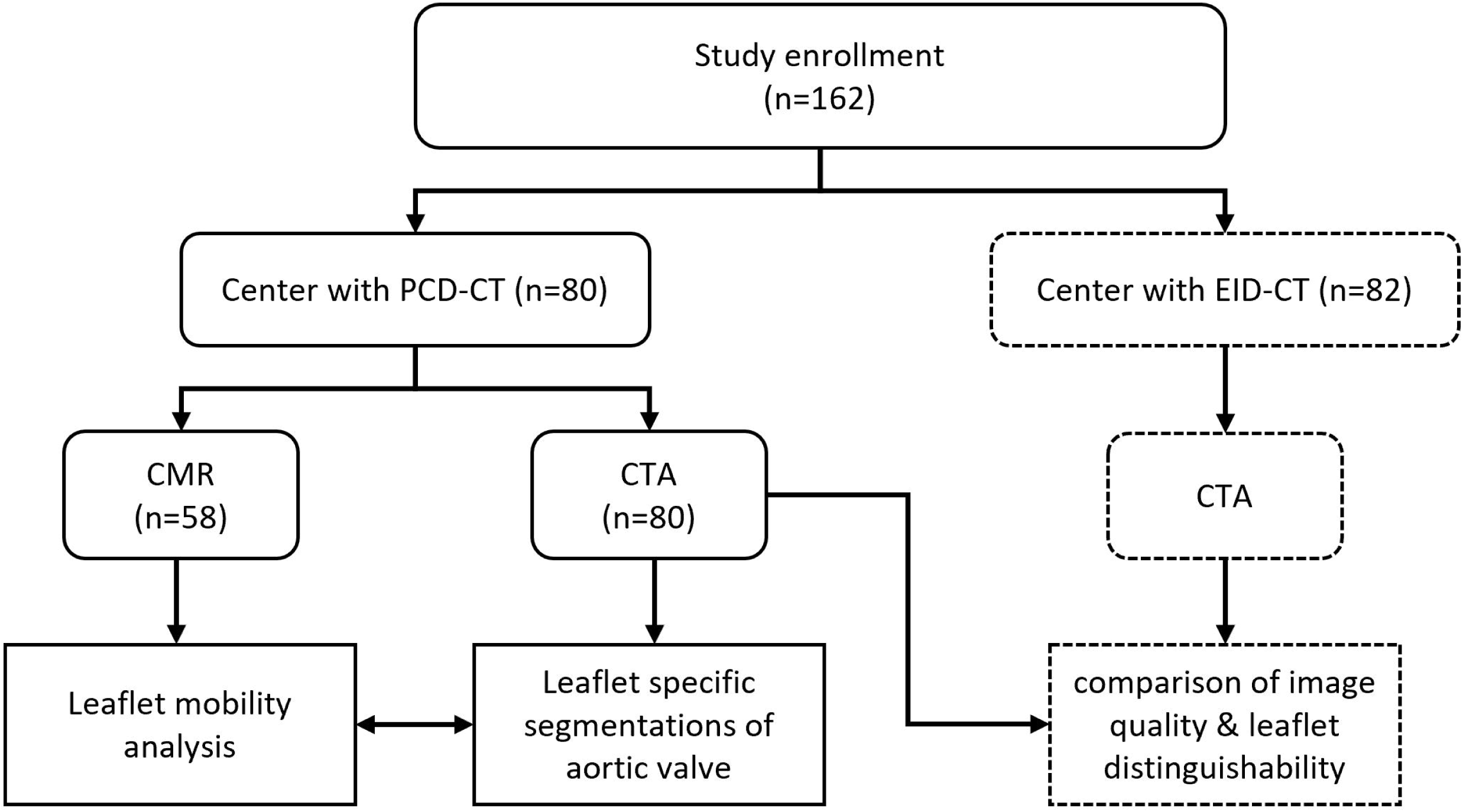
Study flowchart. Participants were enrolled at two centers based on geographic location using the same inclusion criteria and underwent CT angiography (CTA). A subset of participants who underwent photon-counting CT also underwent same-day cardiovascular magnetic resonance (CMR).

### Cardiac CT angiography protocol

All PCD-CT examinations were performed on a dual-source photon-counting detector system (NAEOTOM Alpha, Siemens Healthineers, Forchheim, Germany) using ultra-high-resolution mode. EID-CT scans were acquired using two dual energy scanners (IQon Spectral Detector CT, Philips Healthcare, Best, The Netherlands or Somatom Definition Flash, Siemens Healthineers, Forchheim, Germany). Contrast-enhanced acquisition was uniformly performed with bolus tracking in the ascending aorta. Intravenous iodinated contrast media was injected at 4.5 – 5.5 ml/s (56–70 ml of iohexol [300 mg iodine per ml] followed by 30 ml of saline).

Image acquisition was performed using a prospective ECG-triggered sequence (step-and-shoot) for patients with stable sinus rhythm. Images were reconstructed with a slice thickness of both 0.2 mm and 0.6 for PCD-CT while EID-CT was reconstructed at either 0.6 or 0.75 mm (depending on the scanner) using a sharp kernel. Reconstructions were generated at the best diastolic phase (typically 70–80% of the RR interval). Reconstruction parameters are summarized in supplementary table 4.

### Cardiovascular magnetic resonance

CMR scans were performed on a 1.5 tesla MRI system (Ingenia 1.5 T, Philips Healthcare, The Netherlands). All scans included a gradient echo cine sequence of perpendicular to the aortic valve (“en face” view) (field of view: 250 × 250 mm2, repetition time: 3.1 ms, echo time: 1.54 ms, flip angle: 45°, heart phases: 30, slice thickness: 6 mm, in-plane resolution: 1.89 × 1.96 mm^2^ [reconstructed: 1.04 × 1.04 mm^2^]).

### PCD-CT analysis

Volumetric CT datasets were aligned to the aortic valve plane using multiplanar reformation at the optimal diastolic reconstruction phase following the Society of Cardiac CT (SCCT) guidelines[12]. The aortic annulus was defined at the level of the basal hinge points of the three aortic cusps. Area, average diameter and average HU of blood pool in the aortic annulus were measured. Manual volumetric segmentation of the aortic valve was performed using Slicer 3D (version 5.8.0) and a density-based thresholding approach adapted from Cartlidge et al.[13] (Figure 2A). Tissue components within the valve leaflets were differentiated into calcific volume (defined as hyperattenuating structures within the leaflet) and fibrotic volume (defined as non-calcified leaflet thickening distinct from the blood pool and calcium) (Figure 2A). The lower threshold for detection of calcific volume was 3 SDs above the mean attenuation measured within the blood pool. Careful attention was dedicated to excluding calcifications of the aortic wall. In cases where calcification extended to the anterior mitral leaflet, the annulus defined the anatomical cutoff. A fixed lower threshold of 20 HU was set for fibrotic volumes to exclude photon counting artifacts. Previously, thresholds of 30 and 40 HU have been described for EID-CT[6,13], however we found a lower threshold of 20 HU to be a better fit for PCD-CT, since higher thresholds would exclude valve tissue. The upper threshold of fibrotic volume was set by starting at 200 HU and adjusting by increments of 20 HU in either direction until the whole aortic valve leaflets were segmented without inclusion of blood pool. Fibrotic and calcific volumes were manually assigned to each coronary cusp (right [RCC], left [LCC], non-coronary [NCC]). To correct for variation in valve size among participants, fibrocalcific volumes were indexed to the aortic annulus area as described previously[14]. All segmentations were performed by M. B., repeated segmentations were performed by A. B. for a subset of 10 patients.

**Figure 2.**
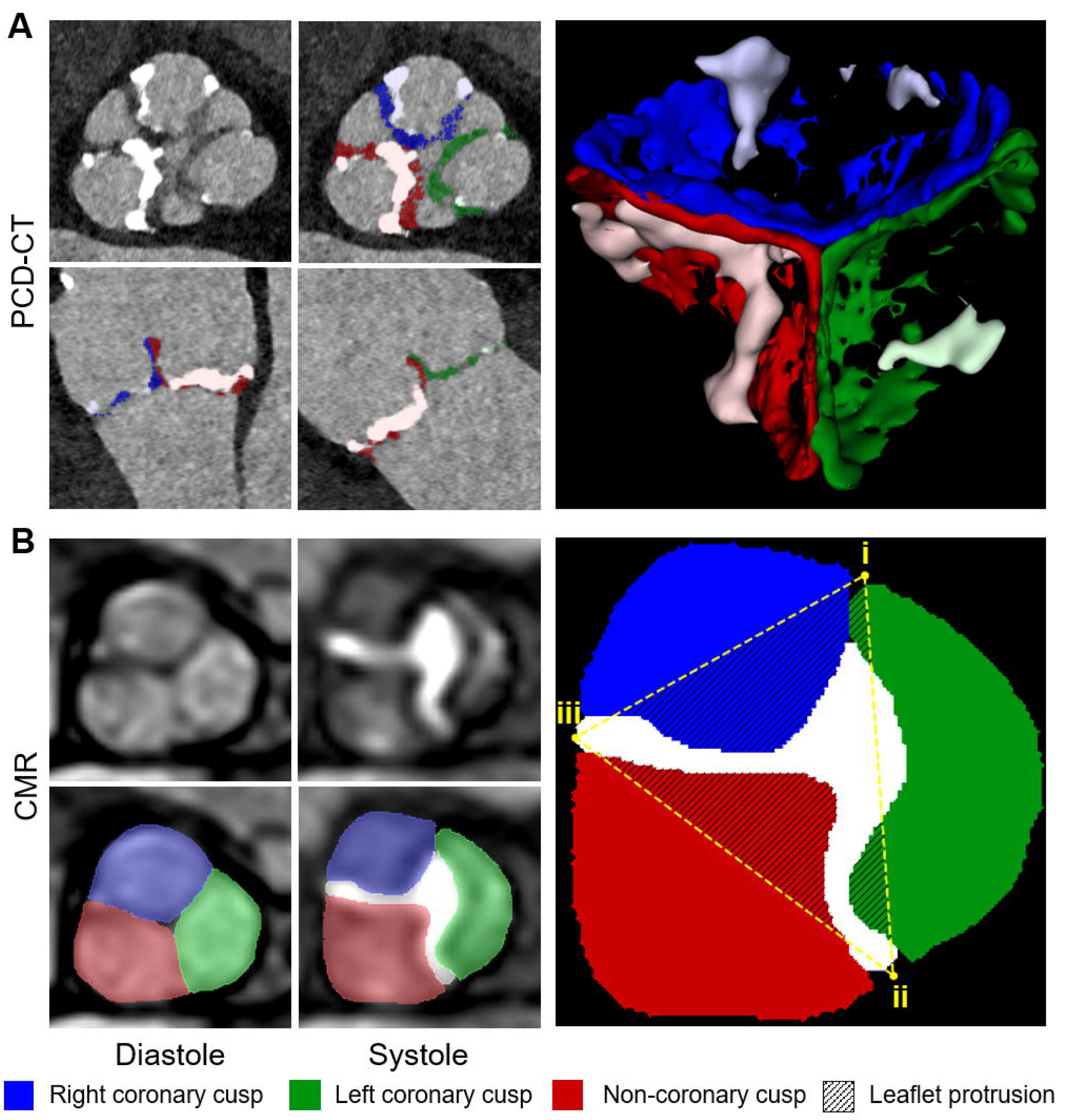
Leaflet segmentations. Representative photon-counting CT (PCD-CT) and CMR images of an aortic valve (male in his 70’s). (A) Ultra-high-resolution PCD-CT images used for manual, slice-by-slice segmentation. Left panel shows in plane (upper row), coronal and sagittal reformations (lower row); solid colors show fibrosis and de-saturated colors show calcification. Right panel shows finished volumetric segmentation of the valve. (B) CMR images with corresponding leaflet segmentation. Upper row: native cine images. Lower row: the same slices with manual segmentations overlaid. Left column: diastole. Middle column: systole. Right image: schematic representation of the segmented valve demonstrating the three leaflets. Yellow points (i-iii) define the commissural meeting points and dashed lines outline the triangular region of interest used for geometric analysis. Hatched areas indicate leaflet protrusion. Notice that the more calcified non-coronary and right coronary leaflets show more protruding valve areas in CMR segmentation while the less calcified left-coronary cusp shows near ideal opening.

### CMR analysis

To derive a leaflet-specific mobility index (LMI), we used a geometric approach using "en face" CMR gradient echo cine images (Figure 2B). Analysis of leaflet excursion was performed in both the end-diastolic to account for different leaflet sizes and peak systolic phases (Figure 1B):

- Three points were defined at the meeting points of three commissural insertions of the aortic valve leaflets: point i between RCC and LCC, point ii between LCC and NCC and point iii between NCC and RCC.
- A triangular ROI was generated by connecting the three meeting points of commissural insertions with a straight line. This triangle ROI serves as an approximated model of the ideal valve orifice at the commissural level.
- The cross-sectional area of leaflet excursion situated within the boundaries of this triangular ROI was segmented manually for each cusp.
- LMI was calculated as the relative reduction in leaflet area within the triangular ROI from diastole (leaflet area_ED_) to systole (leaflet area_PS_). The following formula was applied for each leaflet:

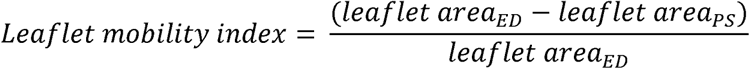
- The mean of the three leaflet mobility indices was also calculated.

A representative illustration of the geometric approach used for leaflet-specific mobility assessment is provided in Supplementary Figure 3. In addition, the planar aortic valve area (AVA_CMR_) was derived from the peak-systolic frame. Repeated segmentation was performed by an additional rater (J.A.L.) for a subset of 30 patients.

### Image quality analysis

Subjective and quantitative image quality of PCD-CT datasets were compared to EID-CT datasets from a different center. Evaluation was performed by two cardiovascular radiologists (J.A.L. with 13 years of experience and M.B. with 2 years of experience), who were blinded to the CT and clinical data. The ability to clearly distinguish all three aortic valve leaflets from each other and adjacent blood pool were evaluated in a binary manner (yes or no). Perceived aortic valve image quality, image noise and aortic root contrast attenuation were rated using a 5-point Likert scale (1=poor, 5=excellent).

Objective image quality was quantified by calculating the signal-to-noise ratio (SNR) and contrast-to-noise ratio (CNR). For SNR, a circular region of interest (ROI) measuring 2 cm was placed in the ascending aorta (between the aortic valve and coronary ostia). CNR was calculated using subcutaneous fat as background reference.

### Statistical analysis

Statistical analysis was performed using R (version 4.6.1, R Foundation for Statistical Computing, Vienna, Austria) and GraphPad Prism (version 10.6.1; GraphPad Software, San Diego, CA).

Continuous variables are presented as mean ± standard deviation (SD) for normally distributed data or median with interquartile range (IQR) for non-normally distributed data. Categorical variables are expressed as frequencies and percentages. Data normality was assessed using the Shapiro-Wilk test prior to statistical analysis. Group comparisons (e.g., differences between sexes) were performed using the Student’s t-test or the Mann-Whitney U test, as appropriate. Fisher’s exact test was used for categorical variables. Spearman’s rank correlation coefficients (ρ) were used for correlation analysis. Bland Altman analysis was used for repeatability assessment of LMI calculation. P <0.05 was considered indicative for statistical significant difference.

## Results

### Patient Cohort

A total of 162 participants were included in this analysis. 80 participants underwent PCD-CT, out of whom 58 also underwent CMR. 82 participants underwent EID-CT at a different center. Baseline clinical characteristics are summarized in Table 1 (PCD-CT) and Table S1 (EID-CT). Most PCD-CT participants presented with moderate AS (57/80 [73%]). Mean AVA_Echo_ was 1.4 ± 0.3 cm^2^ and the mean pressure gradient was 16.6 ± 6.7 mmHg. PCD-CT and EID-CT had similar radiation doses (403.2 ± 215 vs 373.8 ± 169.6 mGycm; p = 0.86).

**Table 1.** Baseline clinical characteristics of the photon-counting CT cohort.

| Variable | Overall<br>n = 80 | Female<br>n = 20 | Male<br>n = 60 | P value |
| --- | --- | --- | --- | --- |
| Age | 72.5 ± 9.5 | 71.9 ± 10.0 | 72.7 ± 9.4 | 0.710 |
| Hypertension | 46 (68%) | 12 (75%) | 34 (65%) | 0.472 |
| Diabetes | 22 (32%) | 7 (44%) | 15 (29%) | 0.265 |
| Hypercholesterolemia | 26 (39%) | 4 (25%) | 22 (43%) | 0.194 |
| Angina | 59 (74%) | 13 (65%) | 46 (77%) | 0.304 |
| Previous myocardial infarction | 9 (13%) | 1 (6.3%) | 8 (16%) | 0.675 |
| Previous coronary artery bypass grafting | 10 (15%) | 1 (6.7%) | 9 (18%) | 0.433 |
| Peak systolic velocity (m/s) | 2.7 ± 0.5 | 2.7 ± 0.5 | 2.7 ± 0.5 | 0.845 |
| Pressure gradient (mmHg) | 16.6 ± 6.7 | 16.8 ± 7.3 | 16.5 ± 6.6 | 0.925 |
| Aortic valve area (cm<sup>2</sup>) | 1.4 ± 0.3 | 1.4 ± 0.3 | 1.4 ± 0.3 | 0.445 |
| Stroke volume index (ml/m<sup>2</sup>) | 37.5 ± 10.9 | 33.1 ± 8.1 | 38.5 ± 11.3 | 0.119 |
| Aortic stenosis severity |  |  |  | 0.569 |
| Mild | 21 (27%) | 4 (21%) | 17 (29%) |  |
| Moderate | 57 (73%) | 15 (79%) | 42 (71%) |  |
| Severe | 0 (0%) | 0 (0%) | 0 (0%) |  |

Data are means ± standard deviation or absolute numbers with percentages in parenthesis.

### Sex-specific analysis of leaflet calcification and fibrosis

Male participants had a larger annular area compared to females (4.92±0.90 cm^2^ vs. 3.63±0.66 cm^2^; p < 0.001) (Table 2). Regarding tissue composition, males demonstrated significantly higher global indexed calcific volume (ICV) compared to females (41.5 ± 33.1 µL/cm^2^ vs. 15 ± 12 µL/cm^2^; p = 0.001), a pattern observed across all three leaflets (e.g. ICV_RCC_: 15.1 ± 16.1 µL/cm^2^ vs. 4.4 ± 6.5 µL/cm^2^; p < 0.001) (Figure 3). In contrast, global indexed fibrotic volume (IFV) was more pronounced in females (140.2 ± 49.0 µL/cm^2^ vs. 104.9 ± 42.9 µL/cm^2^; p = 0.003). Leaflet-specific analysis showed that this pattern was consistent across all aortic leaflets (e.g. LCC: 46.8 ± 17.6 µL/cm^2^ vs. 33.6 ± 15.2 µL/cm^2^). ICV showed high concordance with the non-contrast Agatston score (ρ = 0.91 [0.86, 0.94]; p < 0.001).

**Figure 3.**
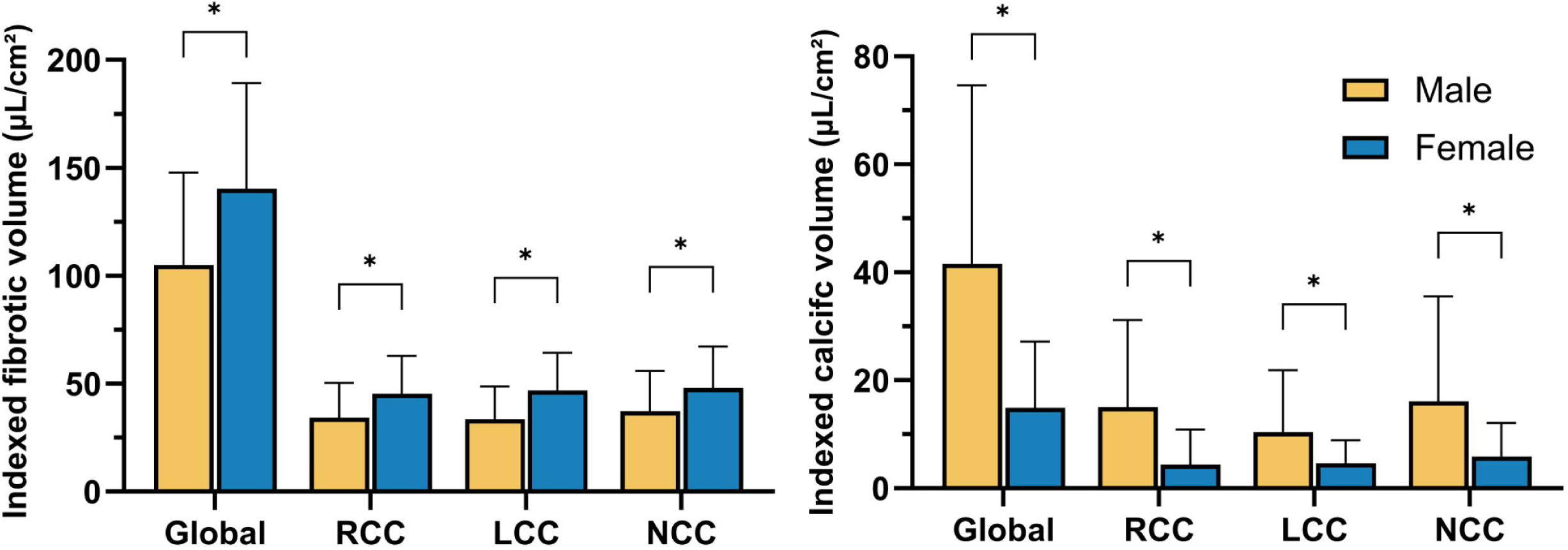
PCD-CT derived distribution of valvular tissue phenotypes. Bar charts showing means of indexed tissue volumes across all leaflets for (A) fibrotic volume and (B) calcific volume. Males (n = 60, yellow) exhibit a predominantly calcific phenotype, while females (n = 16, blue) present with a significantly higher indexed fibrotic burden across all three leaflets. Bars show means; error bars represent standard deviation; * indicates p<0.05.

**Table 2:**
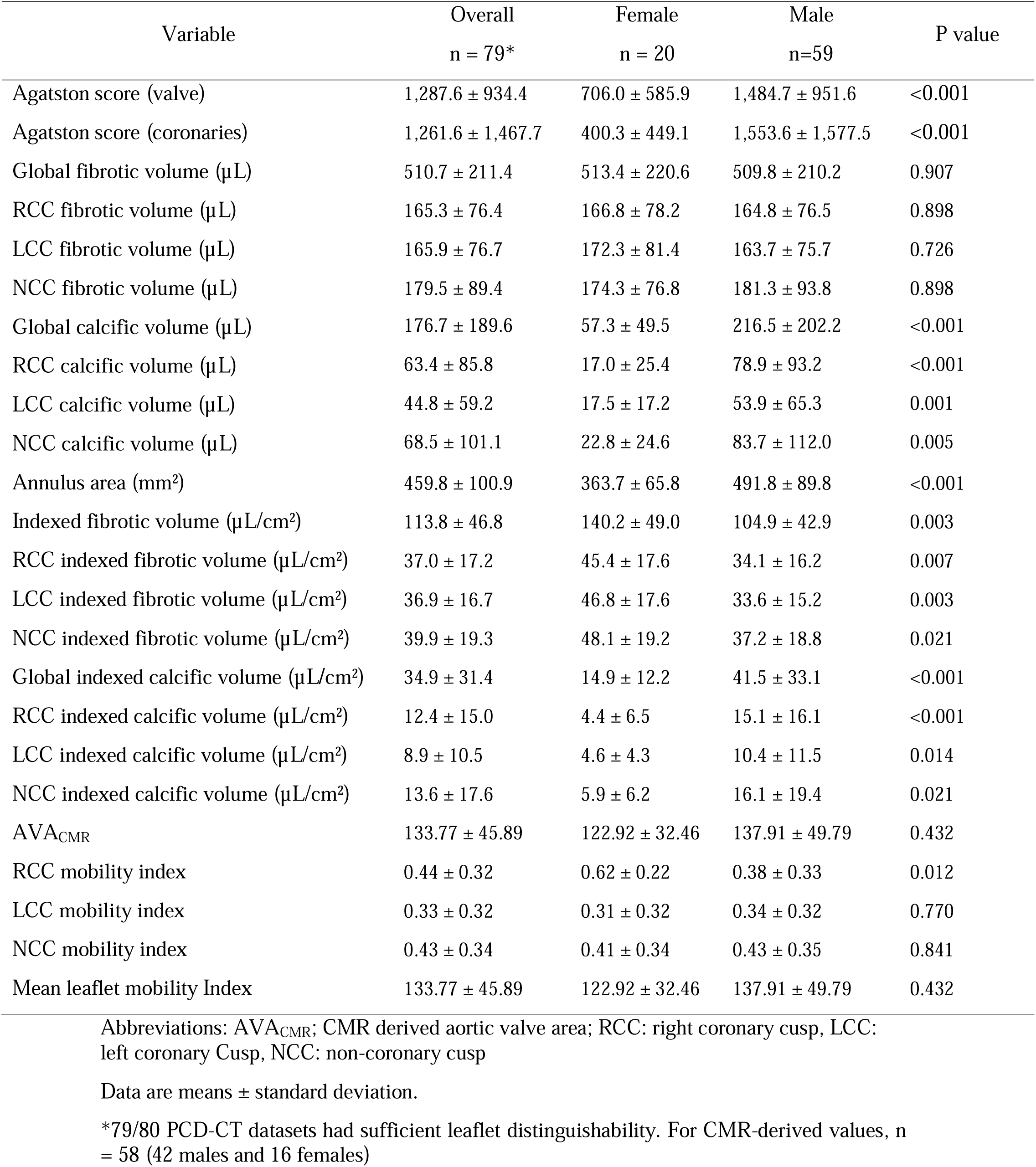
Photon-counting CT and cardiovascular magnetic resonance derived measurements.

| Variable | Overall<br>n = 79* | Female<br>n = 20 | Male<br>n=59 | P value |
| --- | --- | --- | --- | --- |
| Agatston score (valve) | 1,287.6 ± 934.4 | 706.0 ± 585.9 | 1,484.7 ± 951.6 | <0.001 |
| Agatston score (coronaries) | 1,261.6 ± 1,467.7 | 400.3 ± 449.1 | 1,553.6 ± 1,577.5 | <0.001 |
| Global fibrotic volume (μL) | 510.7 ± 211.4 | 513.4 ± 220.6 | 509.8 ± 210.2 | 0.907 |
| RCC fibrotic volume (μL) | 165.3 ± 76.4 | 166.8 ± 78.2 | 164.8 ± 76.5 | 0.898 |
| LCC fibrotic volume (μL) | 165.9 ± 76.7 | 172.3 ± 81.4 | 163.7 ± 75.7 | 0.726 |
| NCC fibrotic volume (μL) | 179.5 ± 89.4 | 174.3 ± 76.8 | 181.3 ± 93.8 | 0.898 |
| Global calcific volume (μL) | 176.7 ± 189.6 | 57.3 ± 49.5 | 216.5 ± 202.2 | <0.001 |
| RCC calcific volume (μL) | 63.4 ± 85.8 | 17.0 ± 25.4 | 78.9 ± 93.2 | <0.001 |
| LCC calcific volume (μL) | 44.8 ± 59.2 | 17.5 ± 17.2 | 53.9 ± 65.3 | 0.001 |
| NCC calcific volume (μL) | 68.5 ± 101.1 | 22.8 ± 24.6 | 83.7 ± 112.0 | 0.005 |
| Annulus area (mm<sup>2</sup>) | 459.8 ± 100.9 | 363.7 ± 65.8 | 491.8 ± 89.8 | <0.001 |
| Indexed fibrotic volume (μL/cm<sup>2</sup>) | 113.8 ± 46.8 | 140.2 ± 49.0 | 104.9 ± 42.9 | 0.003 |
| RCC indexed fibrotic volume (μL/cm<sup>2</sup>) | 37.0 ± 17.2 | 45.4 ± 17.6 | 34.1 ± 16.2 | 0.007 |
| LCC indexed fibrotic volume (μL/cm<sup>2</sup>) | 36.9 ± 16.7 | 46.8 ± 17.6 | 33.6 ± 15.2 | 0.003 |
| NCC indexed fibrotic volume (μL/cm<sup>2</sup>) | 39.9 ± 19.3 | 48.1 ± 19.2 | 37.2 ± 18.8 | 0.021 |
| Global indexed calcific volume (μL/cm<sup>2</sup>) | 34.9 ± 31.4 | 14.9 ± 12.2 | 41.5 ± 33.1 | <0.001 |
| RCC indexed calcific volume (μL/cm<sup>2</sup>) | 12.4 ± 15.0 | 4.4 ± 6.5 | 15.1 ± 16.1 | <0.001 |
| LCC indexed calcific volume (μL/cm<sup>2</sup>) | 8.9 ± 10.5 | 4.6 ± 4.3 | 10.4 ± 11.5 | 0.014 |
| NCC indexed calcific volume (μL/cm<sup>2</sup>) | 13.6 ± 17.6 | 5.9 ± 6.2 | 16.1 ± 19.4 | 0.021 |
| AVA<sub>CMR</sub> | 133.77 ± 45.89 | 122.92 ± 32.46 | 137.91 ± 49.79 | 0.432 |
| RCC mobility index | 0.44 ± 0.32 | 0.62 ± 0.22 | 0.38 ± 0.33 | 0.012 |
| LCC mobility index | 0.33 ± 0.32 | 0.31 ± 0.32 | 0.34 ± 0.32 | 0.770 |
| NCC mobility index | 0.43 ± 0.34 | 0.41 ± 0.34 | 0.43 ± 0.35 | 0.841 |
| Mean leaflet mobility Index | 133.77 ± 45.89 | 122.92 ± 32.46 | 137.91 ± 49.79 | 0.432 |

Abbreviations: AVA<sub>CMR</sub>; CMR derived aortic valve area; RCC: right coronary cusp, LCC: left coronary Cusp, NCC: non-coronary cusp
Data are means ± standard deviation.
\*79/80 PCD-CT datasets had sufficient leaflet distinguishability. For CMR-derived values, n = 58 (42 males and 16 females)

Wilcoxon signed-rank test revealed a higher fibrocalcific ratio of the LCC (53.1 ± 275.9) compared to RCC (19.4 ± 48.1; p < 0.001) and NCC (13.5 ± 34.0; p < 0.001). ICV_LCC_ was lower compared to the ICV_RCC_ (10.4 ± 11.5 vs. 15.1 ± 16.1; p = 0.027) in males and a similar trend for ICV_LCC_ compared to the ICV_NCC_ (16.1 ± 19.4; p = 0.071). ICV_RCC_ and ICV_NCC_ were similar (p =0.615). No differences were observed between individual leaflet ICV in females or IFV in both sexes.

### Leaflet mobility index

A subset of 58 participants (42 males, 16 females) underwent CMR for functional valve assessment. The AVA_CMR_ was similar between males and females (1.38 vs. 1.23 cm^2^, p = 0.5). On a leaflet-level, the RCC was more mobile in females compared to males (LMI_RCC_ = 0.62 ± 0.22 vs. 0.38 ± 0.33; p = 0.012). No significant sex-differences in mobility index were observed for LMI_LCC_ (0.31 ± 0.32 vs. 0.34 ± 0.32; p = 0.77) or the LMI_NCC_ (0.41 ± 0.34 vs. 0.43 ± 0.35; p = 0.432).

### Segmentation data and leaflet mobility index

Spearman’s correlation values are summarized in supplementary Table 2. LMI_RCC_ negatively correlated with ICV_RCC_ (ρ = −0.64 [-0.77, −0.46]; p < 0.001) (Figure 4) but had no significant correlation with IFV_RCC_ (ρ= 0.16 [-0.10, 0.41]; p = 0.219). LMI_NCC_ negatively correlated with ICV_NCC_ (ρ = −0.69 [-0.80, −0.53]; p < 0.001) but had no significant correlation with IFV_NCC_ (ρ = 0.24 [-0.02, 0.47]; p = 0.065). In contrast, LMI_LCC_ negatively correlated with IFV_LCC_ (ρ = - 0.35 [-0.56, −0.10]; p = 0.007) and with ICV_LCC_ (ρ = −0.30 [-0.52, −0.04]; p = 0.023).

**Figure 4.**
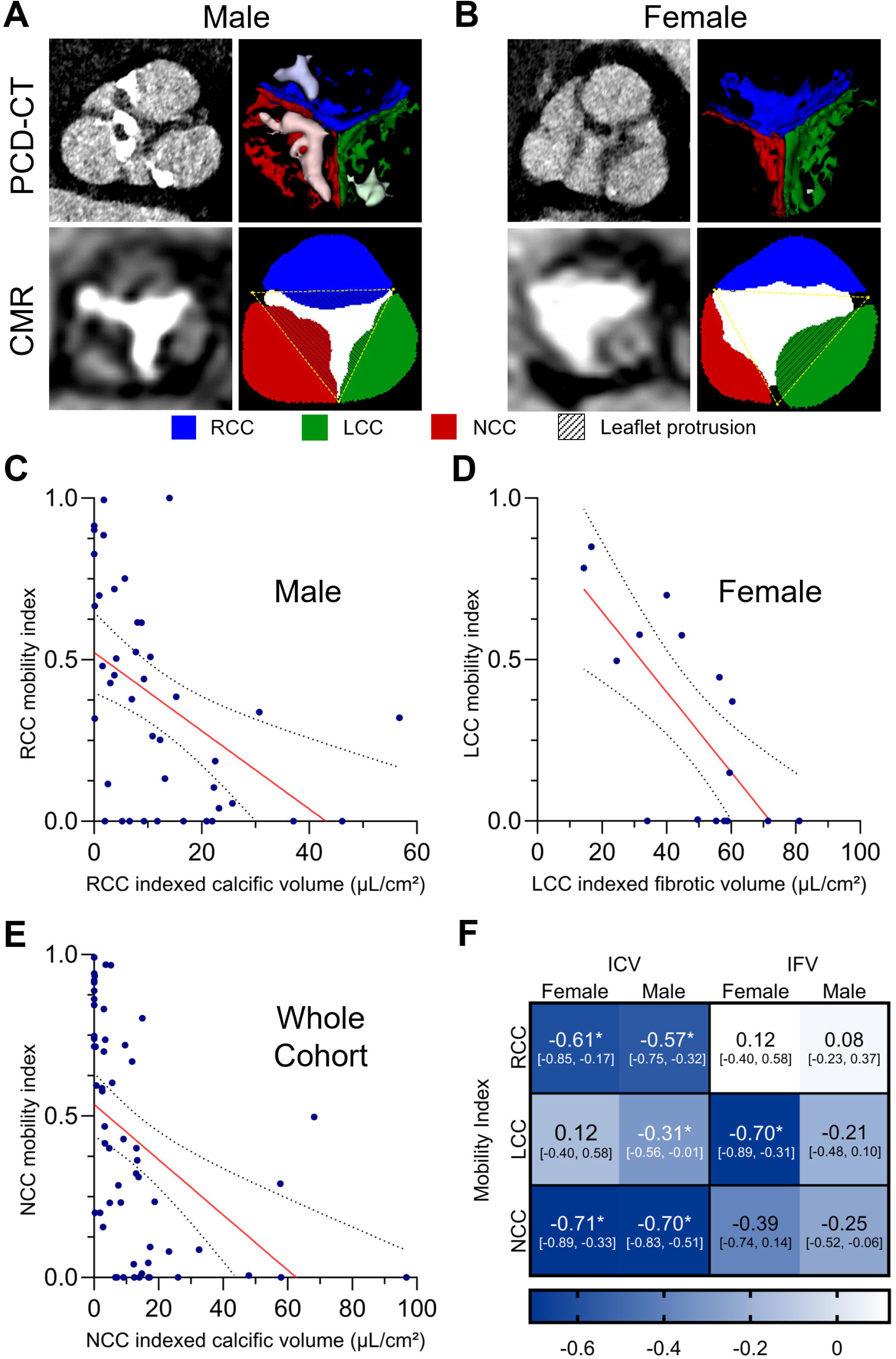
Divergent, sex dependent drivers of leaflet mobility. (A) Representative images of a male in his 70’s. Photon-counting CT (PCD-CT) shows heavy calcification, particularly in the non-coronary cusp (NCC). The corresponding cardiovascular magnetic resonance (CMR) segmentation (bottom) demonstrates that this calcified region coincides with significant leaflet protrusion (hatched area). (B) Representative images of a female in her 70’s. In contrast to the male phenotype, PCD-CT shows fibrosis-dominant thickening without dense calcium. This is most pronounced in the left coronary cusp (LCC), which spatially corresponds to the leaflet protrusion observed in the CMR segmentation. (C–E) Scatter plots correlating leaflet mobility index with indexed tissue volumes. (C) In males, right coronary cusp (RCC) mobility index is inversely correlated with indexed calcific volume (ICV). (D) In females, LCC mobility is significantly driven by indexed fibrotic volume (IFV). (E) NCC mobility index is strongly restricted by ICV across the cohort. (F) Correlation matrix heatmap summarizing the divergent pathophysiology. Values represent Spearman’s rank correlation (ρ). Blue shading indicates a negative correlation (indexed volume vs. mobility index). * indicates p<0.05.

Sex-specific analysis revealed divergent correlations of ICV and IFV with LMI (Figure 4). In male participants, LMI correlated negatively with ICV in all leaflets (e.g. RCC: ρ = −0.57 [- 0.75, −0.32]; p < 0.001), but not with IFV (ρ =0.08 [-0.23, 0.37]; p = 0.630).

In female participants, a negative correlation was observed between LMI_LCC_ and IFV_LCC_ (ρ = −0.70 [-0.89, −0.31]; p = 0.003), but no correlation with ICV_LCC_ (ρ = −0.16 [-0.61, 0.36]; p = 0.550). LMI showed negative correlation with ICV for NCC (ρ −0.71 [-0.89, −0.33]; p = 0.002) and RCC (ρ = −0.61 [-0.85, −0.17]; p = 0.012), but not to IFV of NCC (ρ = −0.39 [0.14, −0.74]; p = 0.140) and RCC (ρ = 0.12 [-0.40, 0.58]; p = 0.648).

### Correlation with clinical data

ICV showed moderate correlation with echocardiographic parameters (Supplementary Figure 1): Positive correlation with peak velocity (Vmax) and mean pressure gradient (MPG) (both ρ = −0.55 [-0.36, −0.69]; p < 0.001) and negative correlation with AVA_Echo_ (ρ = −0.39 [-0.57, - 0.17]; p < 0.001). IFV showed no correlation with echocardiographic parameters. CMR-derived LMI correlated with echocardiographic Vmax (ρ = −0.44 [-0.64, −0.19]; p < 0.001) and MPG (ρ = −0.45 [-0.64, −0.20]; p < 0.001). AVA_CMR_ also correlated negatively with Vmax (ρ = −0.58 [-0.74, −0.37]; p < 0.001) and MPG (ρ = −0.53[-0.71, −0.30]; p < 0.001) while showing weak correlation with AVA_Echo_ (ρ = 0.33 [0.05, 0.56]; p = 0.017).

### Image quality and modality comparison

Qualitative comparison showed superior ratings for PCD-CT regarding overall image quality of the aortic valve (5; IQR [4–5] vs. 3 [3–4]; p<0.001). EID-CT datasets had higher ratings for image noise (5 [4.75–5] vs. 4 [4–4]; p < 0.001). Perceived contrast attenuation of the aortic root received similar ratings (5 [5–5] vs. 5 [5–5]; p=0.268)

Quantitative analysis revealed higher SNR (19.0 ± 7.2 vs. 7.6 ± 3.2; p < 0.001) and CNR (38.17±18.07 vs. 12.12±4.73; p < 0.001) values for EID-CT, primarily attributed to the thinner slice thickness of PCD-CT compared to EID-CT. Aortic valve leaflet distinguishability was higher in PCD-CT compared to EID-CT datasets (79/80 [99%] vs. 42/82 [51%]; p < 0.001) (Figure 5).

**Figure 5.**
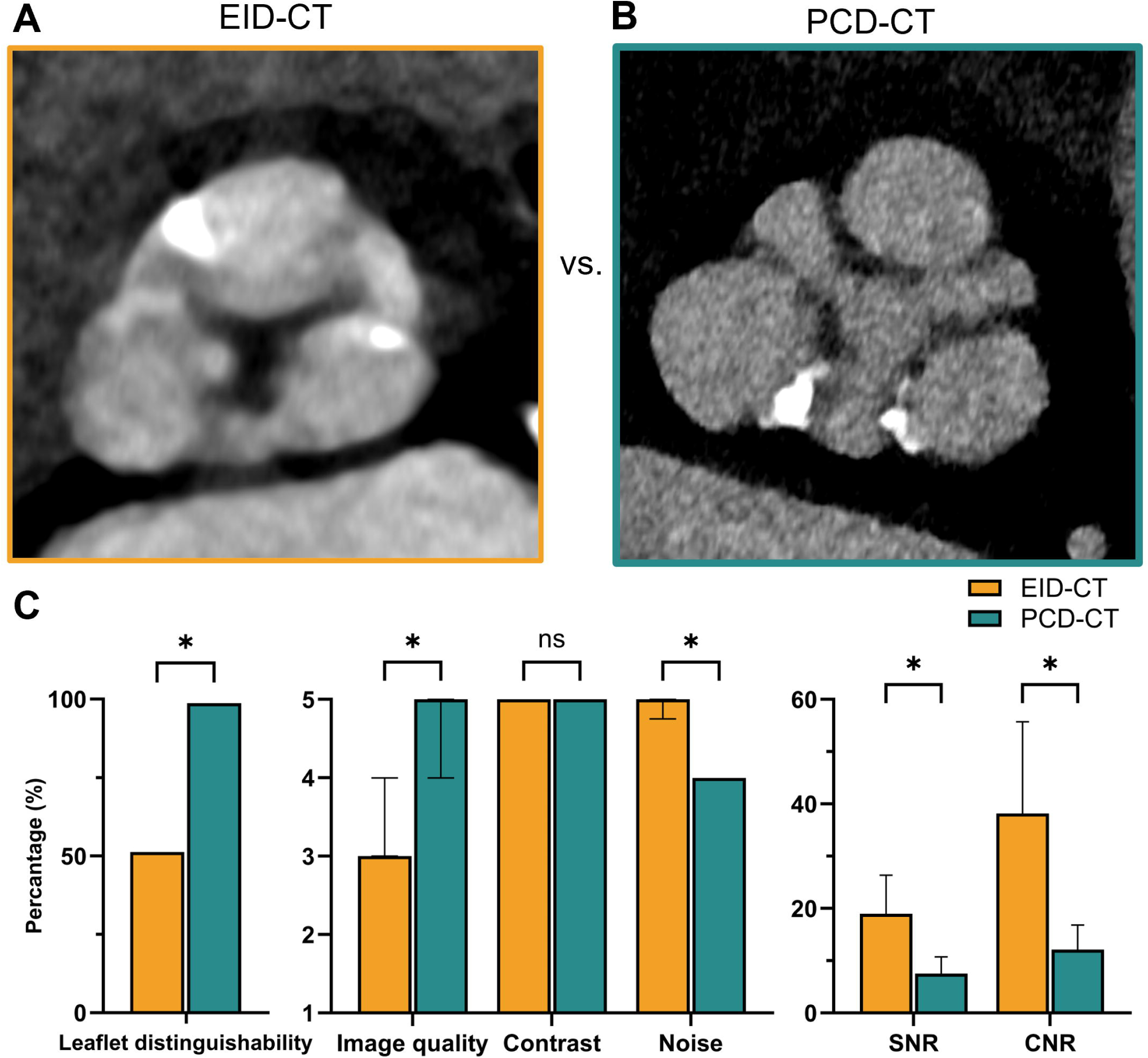
Quality comparison. Representative axial images of the aortic valve from (A) energy-integrating detector CT (EID-CT; n =82) and (B) photon-counting detector CT (PCD-CT; n = 80). PCD-CT shows clear distinguishability of the separate aortic valve leaflets in more participants (76/80; 99%) than EID-CT (42/82; 51%). Despite higher perceived noise, PCD-CT received significantly superior subjective ratings for image quality. Quantitative measurements showed higher signal-to-noise (SNR) and contrast-to-noise (CNR) ratios are in EID-CT. Middle plot show medians with interquartile ranges and right plot shows mean with standard deviation; * indicates p<0.05, ns: not significant.

### Repeatability analysis

Repeated aortic valve segmentations (n=10) showed excellent mean dice score (0.90 ± 0.08) for calcific volumes and high dice scores (0.82 ± 0.9) for fibrotic volumes of individual valve leaflets. Bland-Altman-Analysis (Supplementary Figure 2) demonstrated low bias (5.63 and 18.95) and narrow 95% limits of agreement ([-19.1, 30.3] and [-429.6, 391]) for calcific and fibrotic volumes.

Bland-Altmann analysis of repeated LMI calculations showed strong agreement (mean bias: - 0.009; 95% limits of agreements; −0.093 to 0.076).

## Discussion

This study demonstrates sex-specific differences in aortic stenosis on a per leaflet level: while males predominantly display a calcification pattern, females exhibit a higher burden of valve fibrosis. Furthermore, using a CMR-based mobility index, we showed that these tissue differences translate into distinct functional impairments. Specifically, valve mobility in males is primarily restricted by calcification, whereas in females, fibrotic tissue plays a more pronounced role, particularly in the left coronary leaflet.

The transition from EID-CT to PCD-CT represents a pivotal shift in cardiovascular imaging[15]. While previous studies have established the utility of CT angiography for quantifying the "fibro-calcific" volumes of aortic stenosis[6], EID-CT is limited by blooming artifacts. High-density calcium creates partial volume effects that exaggerate calcific volume and obscure adjacent non-calcified fibrosis[16], which PCD-CT significantly reduces[7,17]. PCD-CT provided superior perceived image quality in our cohort despite the expected increase in image noise associated with thinner slice reconstruction. The improved leaflet distinguishability translated directly into measurable, leaflet-level segmentations not achievable in the majority of our EID-CT datasets.

Recent preliminary work has suggested qualitative benefits of PCD-CT for evaluating native valves in CT angiography[18], however, quantitative segmentations of aortic stenosis using PCD-CT remains absent. The superior leaflet distinguishability in PCD-CT enabled us to transition from global valve assessment to granular, leaflet-specific segmentation. Our data confirms sex-dependent patterns previously described in histopathology[4,19] and standard CTA[6,14]: Males had higher ICV while females exhibited greater IFV, and these differences were consistent across leaflets. We observed a trend toward higher ICVs in the RCC and NCC compared to the LCC in men, which aligns with previous hemodynamic theories regarding flow velocities at the left-ventricular outflow tract[20–22]. Potential biological mechanisms, such as differential expression of extracellular matrix proteins involved in valve maturation[23] or variations in local shear stress profiles, warrant further investigation as drivers of asymmetric valve degeneration.

By segmenting each leaflet separately using the ultra-high resolution mode of PCD-CT, we present an objective quantification of individual leaflet fibrosis, moving beyond previous methods that relied on visual, semi-quantitative ratings of leaflet thickness[20,24]. The ability to non-invasively differentiate between fibrotic and calcific stenosis on a leaflet basis could have significant clinical implications: It would add to our understanding of AS pathophysiology, suggesting that disease progression may vary not only between sexes but also between individual cusps. Furthermore, it may influence therapeutic planning for individualized valve selection and replacement strategy. As calcification is a known predictor of paravalvular leak and device landing zone rupture during TAVI[25,26], precise mapping of calcification versus fibrosis could optimize both patient selection and prosthesis sizing.

A novel aspect of this study is the correlation between tissue composition and individual leaflet mobility derived from same-day CMR. “En-face” sequences of the aortic valve are established as a robust method to measure planimetric AVA[27], with good correlation to TTE derived AVA in severe AS[28,29]. In our cohort AVA_CMR_ showed moderate correlation to AVA_Echo_. This may be due to most of our participants exhibiting only mild and moderate AS, where AVA_Echo_ tends to be more variable than peak systolic velocity and gradient[30]. Consistent with prior work by Chen et al.[20] analyzing systolic excursion using CT, CMR-derived leaflet mobility of the RCC and NCC was primarily associated with calcific burden, whereas LCC mobility, particularly in females, appeared to be more strongly influenced by fibrosis. Future studies could investigate how the localization of fibrocalcific volumes within the leaflets (e.g., annular versus free-edge) influences valve mechanics and functional impairment[31].

This study has some limitations. Only 16 female participants were evaluated in the mobility analysis and results should be interpreted with caution. Additionally, the EID-CT cohort included some patients with severe stenosis (15%) in contrast to no patients within the PCD-CT cohort, potentially contributing to reduced leaflet distinction. Finally, longitudinal follow-up of the C06 cohort is required to determine whether PCD-CT-derived leaflet-specific fibrocalcific volumes are predictive of disease progression.

## Conclusion

Our exploratory study utilized the superior spatial resolution of ultra-high resolution PCD-CT to quantify the fibrotic and calcific burden of each leaflet in participants with moderate aortic stenosis. We observed sex-specific differences across all leaflets and demonstrated how fibrocalcific volumes differently affect CMR-derived individual leaflet mobility. More detailed segmentation using PCD-CT technology may provide further insights into aortic valve morphology in aortic stenosis in the future.

## Data Availability

All data produced in the present study are available upon reasonable request to the authors.

## Acknowledgments

None.

## Data Statement

Data generated or analyzed during the study are available from the corresponding author upon reasonable request.

## Abbreviations

AS: Aortic stenosis
AVA: Aortic valve area
CMR: Cardiovascular magnetic resonance
CNR: Contrast-to-noise ratio
CT: Computed tomography
EID: Energy-integrating detector
ICV: Indexed calcific volume
IFV: Indexed fibrotic volume
IQR: Interquartile range
LMI: Leaflet mobility index
LCC: Left coronary cusp
NCC: Non-coronary cusp
PCD: Photon-counting detector
RCC: Right coronary cusp
ROI: Region of interest
SNR: Signal-to-noise ratio

## References

[1] R.L.J. Osnabrugge, D. Mylotte, S.J. Head, N.M. Van Mieghem, V.T. Nkomo, C.M. LeReun, A.J.J.C. Bogers, N. Piazza, A.P. Kappetein, Aortic Stenosis in the Elderly, JACC 62 (2013) 1002–1012. 10.1016/j.jacc.2013.05.015.

[2] M.-A. Clavel, J. Malouf, D. Messika-Zeitoun, P.A. Araoz, H.I. Michelena, M. Enriquez-Sarano, Aortic Valve Area Calculation in Aortic Stenosis by CT and Doppler Echocardiography, JACC Cardiovasc. Imaging 8 (2015) 248–257. 10.1016/j.jcmg.2015.01.009.

[3] P.R. Goody, M.R. Hosen, D. Christmann, S.T. Niepmann, A. Zietzer, M. Adam, F. Bönner, S. Zimmer, G. Nickenig, F. Jansen, Aortic Valve Stenosis, Arterioscler. Thromb. Vasc. Biol. 40 (2020) 885–900. 10.1161/ATVBAHA.119.313067.

[4] L. Simard, N. Côté, F. Dagenais, P. Mathieu, C. Couture, S. Trahan, Y. Bossé, S. Mohammadi, S. Pagé, P. Joubert, M.-A. Clavel, Sex-Related Discordance Between Aortic Valve Calcification and Hemodynamic Severity of Aortic Stenosis, Circ. Res. 120 (2017) 681–691. 10.1161/CIRCRESAHA.116.309306.

[5] T. Pawade, M.-A. Clavel, C. Tribouilloy, J. Dreyfus, T. Mathieu, L. Tastet, C. Renard, M. Gun, W.S.A. Jenkins, L. Macron, J.W. Sechrist, J.M. Lacomis, V. Nguyen, L. Galian Gay, H. Cuéllar Calabria, I. Ntalas, T.R.G. Cartlidge, B. Prendergast, R. Rajani, A. Evangelista, J.L. Cavalcante, D.E. Newby, P. Pibarot, D. Messika Zeitoun, M.R. Dweck, Computed Tomography Aortic Valve Calcium Scoring in Patients With Aortic Stenosis, Circ. Cardiovasc. Imaging 11 (2018) e007146. 10.1161/CIRCIMAGING.117.007146.

[6] M. Lembo, S.S. Joshi, J. Geers, R. Bing, L. Carnevale, T.A. Pawade, M.K. Doris, E. Tzolos, K. Grodecki, S. Cadet, N. Craig, T. Singh, P.J. Slomka, A. White, A. Guala, J.F. Rodriguez-Palomares, A. Ruiz-Muñoz, L. Dux-Santoy, G. Teixido-Tura, L. Galian-Gay, M.C. Williams, D.E. Newby, S. Kwak, S.-P. Lee, A. Powers, M.-A. Clavel, D. Dey, M.R. Dweck, Quantitative Computed Tomography Angiography for the Evaluation of Valvular Fibrocalcific Volume in Aortic Stenosis, JACC Cardiovasc. Imaging 17 (2024) 1351–1362. 10.1016/j.jcmg.2024.06.007.

[7] S.A. Si-Mohamed, S. Boccalini, H. Lacombe, A. Diaw, M. Varasteh, P.-A. Rodesch, R. Dessouky, M. Villien, V. Tatard-Leitman, T. Bochaton, P. Coulon, Y. Yagil, E. Lahoud, K. Erhard, B. Riche, E. Bonnefoy, G. Rioufol, G. Finet, C. Bergerot, L. Boussel, J. Greffier, P.C. Douek, Coronary CT Angiography with Photon-counting CT: First-In-Human Results, Radiology 303 (2022) 303–313. 10.1148/radiol.211780.

[8] M. Boussoussou, M. Vecsey-Nagy, Z. Jokkel, B. Vattay, A. Kubovje, B. Sipos, M. Kolossváry, A.I. Nagy, L. Száraz, S. Beke, B. Schmidt, M. Kiss, B. Merkely, J.A. Decker, T. Emrich, A. Varga-Szemes, P. Maurovich-Horvat, B. Szilveszter, Comparative analysis of photon-counting and energy-integrating detector CT to identify obstructive coronary artery disease, Eur. Radiol. 36 (2026) 3091–3102. 10.1007/s00330-025-12118-7.

[9] N. Willner, G. Prosperi-Porta, L. Lau, A.Y. Nam Fu, K. Boczar, A. Poulin, P. Di Santo, R.R. Unni, S. Visintini, P.E. Ronksley, K.-L. Chan, L. Beauchesne, I.G. Burwash, D. Messika-Zeitoun, Aortic Stenosis Progression, JACC Cardiovasc. Imaging 16 (2023) 314–328. 10.1016/j.jcmg.2022.10.009.

[10] Y. Bohbot, C. Renard, A. Manrique, F. Levy, S. Maréchaux, B.L. Gerber, C. Tribouilloy, Usefulness of Cardiac Magnetic Resonance Imaging in Aortic Stenosis, Circ. Cardiovasc. Imaging 13 (2020) e010356. 10.1161/CIRCIMAGING.119.010356.

[11] K. Woldendorp, P.G. Bannon, S.M. Grieve, Evaluation of aortic stenosis using cardiovascular magnetic resonance: a systematic review & meta-analysis, J. Cardiovasc. Magn. Reson. 22 (2020) 45. 10.1186/s12968-020-00633-z.

[12] P. Blanke, J.R. Weir-McCall, S. Achenbach, V. Delgado, J. Hausleiter, H. Jilaihawi, M. Marwan, B.L. Nørgaard, N. Piazza, P. Schoenhagen, J.A. Leipsic, Computed Tomography Imaging in the Context of Transcatheter Aortic Valve Implantation (TAVI)/Transcatheter Aortic Valve Replacement (TAVR), JACC Cardiovasc. Imaging 12 (2019) 1–24. 10.1016/j.jcmg.2018.12.003.

[13] T.R. Cartlidge, R. Bing, J. Kwiecinski, E. Guzzetti, T.A. Pawade, M.K. Doris, P.D. Adamson, D. Massera, M. Lembo, F.E.C.M. Peeters, C. Couture, D.S. Berman, D. Dey, P. Slomka, P. Pibarot, D.E. Newby, M.-A. Clavel, M.R. Dweck, Contrast-enhanced computed tomography assessment of aortic stenosis, Heart 107 (2021) 1905–1911. 10.1136/heartjnl-2020-318556.

[14] J. Geers, N. Craig, K. Grodecki, M. Lembo, S.S. Joshi, T. Singh, R. Bing, J. Kwieciński, L. Carnevale, D. Kimenai, S. Kwak, S.-P. Lee, K. Patel, T. Treibel, A.R. Muñoz, J.F. Rodriguez Palomares, J.-K. Song, M.-A. Clavel, P. Piña, D. Lorenzatti, A. Latib, L. Slipczuk, R. Rajani, V. Androshchuk, J. Niemierko, M. Grygier, A. Walczak, D. Jagielak, M. Protasiewicz, W. Wojakowski, J. Kochman, Z. Huczek, N.S.B. Mogensen, J.S. Dahl, V. Patel, T. Garg, H. Jilaihawi, R. Makkar, S. Droogmans, B. Cosyns, P. Slomka, M.C. Williams, D.E. Newby, D. Dey, M.R. Dweck, Indexed Aortic Valve Calcium Volume by Computed Tomography Angiography in Patients With Aortic Stenosis: Results of an International Multicenter Cohort Study, JACC Cardiovasc. Imaging (2025). 10.1016/j.jcmg.2025.09.013.

[15] S.P. Sharma, M.-J.D.K. Lemmens, M.W. Smulders, R.P.J. Budde, A. Hirsch, C. Mihl, Photon-counting detector computed tomography in cardiac imaging, Neth. Heart J. Mon. J. Neth. Soc. Cardiol. Neth. Heart Found. 32 (2024) 405–416. 10.1007/s12471-024-01904-5.

[16] M. Sandstedt, J. Marsh, K. Rajendran, H. Gong, S. Tao, A. Persson, S. Leng, C. McCollough, Improved coronary calcification quantification using photon-counting-detector CT: an ex vivo study in cadaveric specimens, Eur. Radiol. 31 (2021) 6621–6630. 10.1007/s00330-021-07780-6.

[17] J.R. Rajagopal, F. Farhadi, T. Richards, M. Nikpanah, P. Sahbaee, S.M. Shanbhag, W.P. Bandettini, B. Saboury, A.A. Malayeri, W.F. Pritchard, E.C. Jones, E. Samei, M.Y. Chen, Evaluation of Coronary Plaques and Stents with Conventional and Photon-counting CT: Benefits of High-Resolution Photon-counting CT, Radiol. Cardiothorac. Imaging 3 (2021) e210102. 10.1148/ryct.2021210102.

[18] C. Mayard, S. Si-Mohamed, A. Houmeau, C. Prieur, J.N. Dacher, L. Boussel, P. Douek, S. Boccalini, Benefits of photon counting CT for the assessment of native heart valves, Eur. J. Radiol. 0 (2026). 10.1016/j.ejrad.2026.112661.

[19] K. Grodecki, A. Olasińska-Wiśniewska, A. Cyran, T. Urbanowicz, J. Kwieciński, J. Geers, B.K. Tamarappoo, B. Perek, R. Gocoł, J. Nawara-Skipirzepa, M. Jemielity, J. Kochman, W. Wojakowski, B. Górnicka, P.J. Slomka, H. Jilaihawi, R.R. Makkar, Z. Huczek, D. Dey, Quantification of Aortic Valve Fibrotic and Calcific Tissue from CTA: Prospective Comparison with Histology, Radiology 312 (2024) e240229. 10.1148/radiol.240229.

[20] I.Y. Chen, V. Vedula, S.B. Malik, T. Liang, A.Y. Chang, K.S. Chung, N. Sayed, P.S. Tsao, J.C. Giacomini, A.L. Marsden, J.C. Wu, Preoperative Computed Tomography Angiography Reveals Leaflet-Specific Calcification and Excursion Patterns in Aortic Stenosis, Circ. Cardiovasc. Imaging 14 (2021) 1122–1132. 10.1161/CIRCIMAGING.121.012884.

[21] M. Kupari, P. Hekali, V.P. Poutanen, Cross sectional profiles of systolic flow velocities in left ventricular outflow tract of normal subjects., Heart 74 (1995) 34–39. 10.1136/hrt.74.1.34.

[22] Calcific Aortic Stenosis: A Disease of the Valve and the Myocardium | JACC, (n.d.). https://www.jacc.org/doi/abs/10.1016/j.jacc.2012.02.093 (accessed February 2, 2026).

[23] Y. Lin, Q. Yang, X. Lin, X. Liu, Y. Qian, D. Xu, N. Cao, X. Han, Y. Zhu, W. Hu, X. He, Z. Yu, X. Kong, L. Zhu, Z. Zhong, K. Liu, B. Zhou, Y. Wang, J. Peng, W. Zhu, J. Wang, Extracellular Matrix Disorganization Caused by ADAMTS16 Deficiency Leads to Bicuspid Aortic Valve With Raphe Formation, Circulation 149 (2024) 605–626. 10.1161/CIRCULATIONAHA.123.065458.

[24] O. Nemchyna, S. Soltani, N. Solowjowa, F. Schoenrath, Y. Hrytsyna, A. Unbehaun, J. Kempfert, J. Stein, C. Knosalla, A. Hagendorff, F. Knebel, V. Falk, J. Knierim, Validity of visual assessment of aortic valve morphology in patients with aortic stenosis using two-dimensional echocardiography, Int. J. Cardiovasc. Imaging 37 (2021) 813–823. 10.1007/s10554-020-02048-4.

[25] S. Milhorini Pio, J. Bax, V. Delgado, How valvular calcification can affect the outcomes of transcatheter aortic valve implantation, Expert Rev. Med. Devices 17 (2020) 773–784. 10.1080/17434440.2020.1789456.

[26] B. Fujita, M. Kütting, M. Seiffert, S. Scholtz, S. Egron, E. Prashovikj, J. Börgermann, T. Schäfer, W. Scholtz, R. Preuss, J. Gummert, U. Steinseifer, S.M. Ensminger, Calcium distribution patterns of the aortic valve as a risk factor for the need of permanent pacemaker implantation after transcatheter aortic valve implantation, Eur. Heart J. - Cardiovasc. Imaging 17 (2016) 1385–1393. 10.1093/ehjci/jev343.

[27] J.L. Cavalcante, O.O. Lalude, P. Schoenhagen, S. Lerakis, Cardiovascular Magnetic Resonance Imaging for Structural and Valvular Heart Disease Interventions, JACC Cardiovasc. Interv. 9 (2016) 399–425. 10.1016/j.jcin.2015.11.031.

[28] C. Mantini, G. Di Giammarco, J. Pizzicannella, S. Gallina, F. Ricci, E. D’Ugo, M. Marchetti, A.R. Cotroneo, N. Ahmed, C. Bucciarelli-Ducci, A. Tartaro, R. De Caterina, Grading of aortic stenosis severity: a head-to-head comparison between cardiac magnetic resonance imaging and echocardiography, Radiol. Med. (Torino) 123 (2018) 643–654. 10.1007/s11547-018-0895-2.

[29] S. Wong, R. Spina, S. Toemoe, K. Dhital, Is cardiac magnetic resonance imaging as accurate as echocardiography in the assessment of aortic valve stenosis?, Interact. Cardiovasc. Thorac. Surg. 22 (2016) 480–486. 10.1093/icvts/ivv362.

[30] D. Manna, M. Eliasson, O. Bech-Hanssen, T. Lindow, Reproducibility of Echocardiographic Measures of Aortic Stenosis Severity and its Impact on Grading of Severity, J. Am. Soc. Echocardiogr. 37 (2024) 370–372.e2. 10.1016/j.echo.2023.11.006.

[31] C. Gollmann-Tepeköylü, F. Nägele, C. Engler, L. Stoessel, B. Zellmer, M. Graber, J. Hirsch, L. Pölzl, E. Ruttmann, I. Tancevski, C. Tiller, F. Barbieri, L. Stastny, S.J. Reinstadler, U.C. Oezpeker, S. Semsroth, N. Bonaros, M. Grimm, G. Feuchtner, J. Holfeld, Different calcification patterns of tricuspid and bicuspid aortic valves and their clinical impact, Interact. Cardiovasc. Thorac. Surg. 35 (2022) ivac274. 10.1093/icvts/ivac274.

