## Supplementary material for "Ultra-high-resolution Photon-Counting CT for Leaflet-Specific Evaluation of Fibrosis and Calcification in Aortic Stenosis": SupplementaryMaterial_clean.docx

**Supplementary Figure 1:**

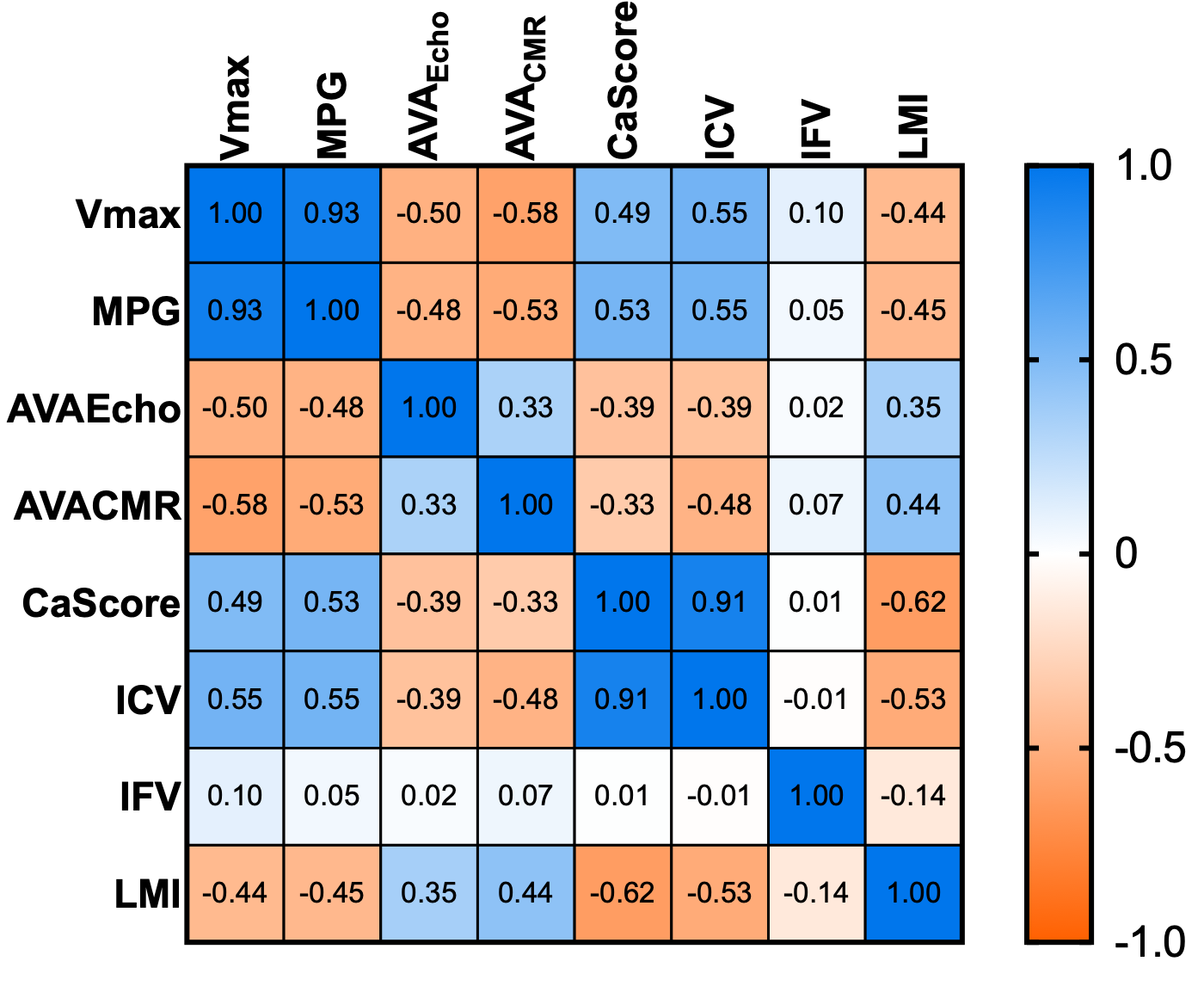

**Correlation matrix of imaging parameters.** Spearman rank correlation values of different imaging modalities of the aortic valve. 95%CI and p values are provided in supplementary table 3.

Abbreviations: Vmax: peak velocity; MPG: mean pressure gradient; AVA: aortic valve area; Echo: Echocardiography; CMR: cardiovascular magnetic resonance; CaScore: Agatston calcium scoring; ICV: indexed calcific volume; IFV: indexed fibrotic volume; LMI: leaflet mobility index.

**Supplementary Figure 2:**

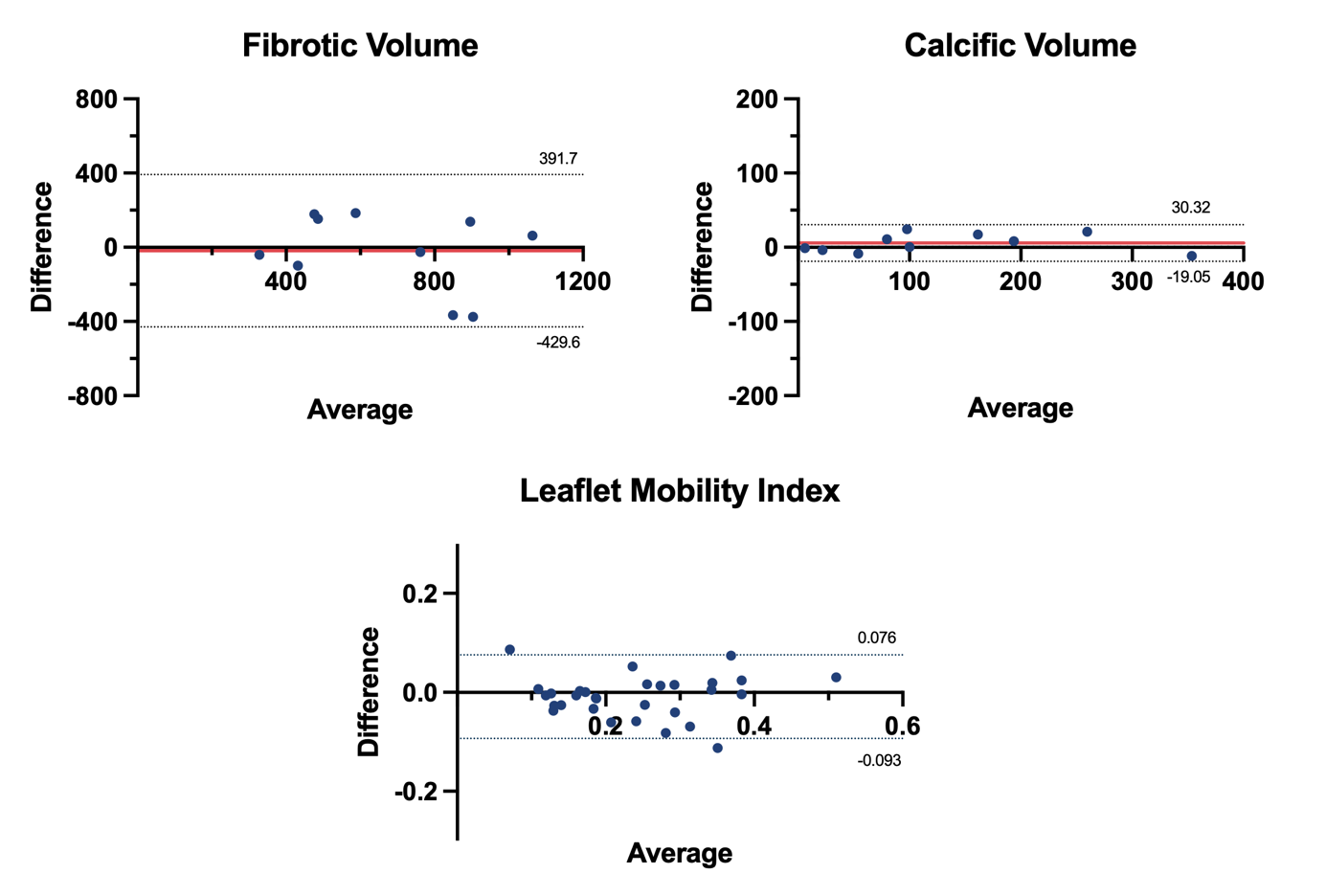

**Segmentation repeatability.** Bland Altman plot of repeated PCD-CT segmentation values and CMR-derived leaflet mobility index (LMI) measurements shows narrow 95% limits of agreement (dotted lines) and low bias (red lines).

**Supplementary Figure 3:**

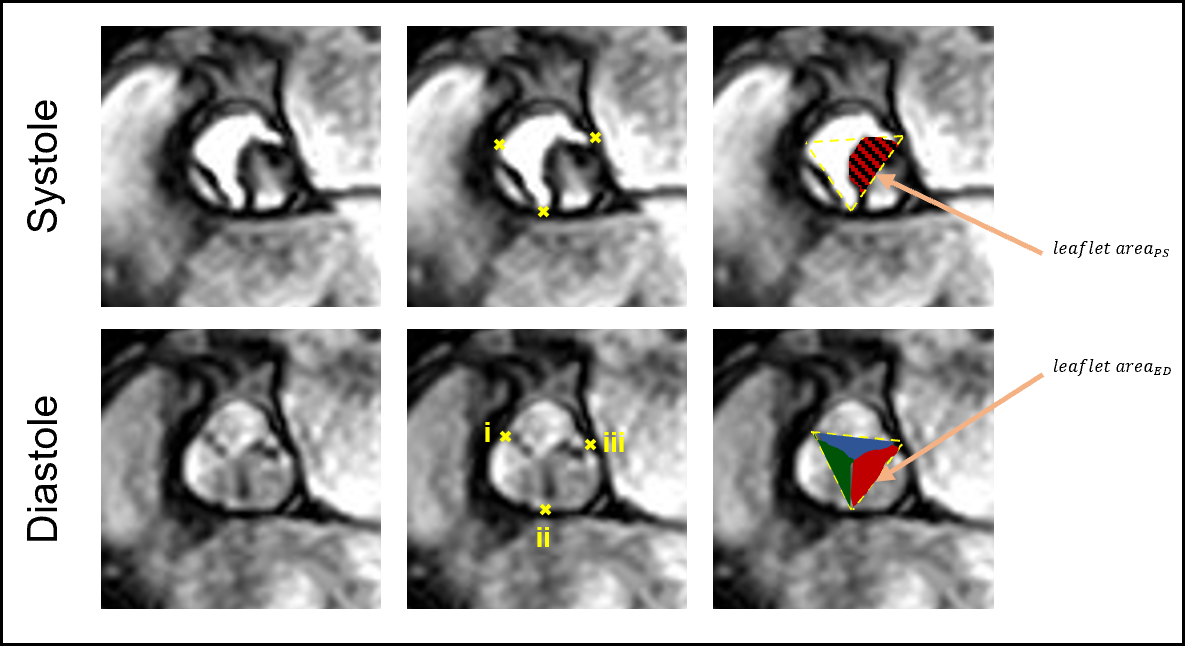

**Geometric assessment of leaflet mobility using en face CMR.** Representative end-diastolic (bottom row) and peak-systolic (top row) en face CMR gradient-echo cine images illustrating the geometric approach used to derive the leaflet-specific mobility index (LMI). In the middle panels, the three commissural insertion points are identified (i: between the right coronary cusp [RCC] and left coronary cusp [LCC]; ii: between the LCC and non-coronary cusp [NCC]; iii: between the NCC and RCC). The three points are connected to define a triangular region of interest, representing an approximation of the ideal valve orifice at the commissural level. In the right panels, the leaflet area within the triangular ROI is manually delineated for each cusp at end-diastole (leaflet area_ED_) and peak systole (leaflet area_PS_). The relative reduction in leaflet area from end-diastole to peak systole was used to calculate the leaflet-specific mobility index. Notice the reduced mobility of the NCC compared to the RCC and LCC.

**Supplementary Table 1:** Baseline clinical characteristics of the energy-integrating detector CT cohort.

| Variable | Overall n = 82 | Female n = 22 | Male n = 60 | P value |
| --- | --- | --- | --- | --- |
| Age | 79.6 ± 7.9 | 78.2 ± 6.9 | 80.1 ± 8.3 | 0.218 |
| Hypertension | 66 (80%) | 15 (68%) | 51 (85%) | 0.117 |
| Diabetes | 21 (26%) | 5 (23%) | 16 (27%) | 0.717 |
| Hypercholesterolemia | 47 (57%) | 11 (50%) | 36 (60%) | 0.417 |
| Angina | 73 (89%) | 73 (89%) | 54 (90%) | 0.696 |
| Previous myocardial infarction | 17 (21%) | 2 (9.1%) | 15 (25%) | 0.137 |
| Previous coronary artery bypass grafting | 12 (15%) | 1 (4.5%) | 11 (18%) | 0.166 |
| Peak systolic velocity (m/s) | 2.8 ± 0.5 | 3.0 ± 0.5 | 2.7 ± 0.4 | 0.019 |
| Pressure gradient (mmHg) | 18.7 ± 6.6 | 21.9 ± 7.2 | 17.4 ± 6.0 | 0.017 |
| Aortic valve area (cm²) | 1.3 ± 0.4 | 1.2 ± 0.6 | 1.3 ± 0.3 | <0.001 |
| Stroke volume index (ml/m²) | 39.8 ± 10.2 | 41.4 ± 11.7 | 39.1 ± 9.6 | 0.558 |
| Aortic stenosis severity |  |  |  | <0.001 |
| Mild | 10 (13%) | 1 (4.5%) | 9 (16%) |  |
| Moderate | 56 (72%) | 12 (55%) | 44 (79%) |  |
| Severe | 12 (15%) | 9 (41%) | 3 (5.4%) |  |

Data are means ± standard deviation or absolute numbers with percentages in parenthesis.

**Supplementary Table 2:** Spearman’s rank correlation of PCD-CT and CMR derived measurements

| Correlation of indexed volume and leaflet mobility index | Overall n = 58 | Female n = 16 | Male n = 42 |
| --- | --- | --- | --- |
| LMI_RCC_ and ICV_RCC_ | -0.64 [-0.77, -0.46] (p < 0.001) | -0.61 [-0.85, -0.17] (p = 0.012) | -0.57 [-0.75, -0.32] (p < 0.001) |
| LMI_RCC_ and IFV_RCC_ | 0.16 [-0.10, 0.41] (p = 0.219) | 0.12 [-0.40, 0.58] (p = 0.648) | 0.08 [-0.23, 0.37] (p = 0.630) |
| LMI_LCC_ and ICV_LCC_ | -0.30 [-0.52, -0.04] (p = 0.023) | -0.16 [-0.61, 0.36] (p = 0.550) | -0.31 [-0.56, -0.01] (p = 0.044) |
| LMI_LCC_ and IFV_LCC_ | -0.35 [-0.56, -0.10] (p = 0.007) | -0.70 [-0.89, -0.31] (p = 0.003) | -0.21 [-0.48, 0.10] (p = 0.190) |
| LMI_NCC_ and ICV_NCC_ | -0.69 [-0.80, -0.53] (p < 0.001) | -0.71 [-0.89, -0.33] (p = 0.002) | -0.70 [-0.83, -0.51] (p < 0.001) |
| LMI_NCC_ and IFV_NCC_ | 0.24 [-0.02, 0.47] (p = 0.065) | -0.39 [0.14, -0.74] (p = 0.140) | -0.25 [- 0.52, -0.06] (p = 0.110) |

Abbreviations: LMI: leaflet mobility index; ICV: indexed calcific volume; IFV: indexed fibrotic volume; RCC: right coronary cusp, LCC: left coronary cusp, NCC: non-coronary cusp

**Supplementary Table 3:** Correlation values of imaging parameters.

|  | Vmax | MPG | AVA_Echo_ | AVA_CMR_ | CaScore | ICV | IFV | LMI |
| --- | --- | --- | --- | --- | --- | --- | --- | --- |
| Vmax | — | 0.93 [0.89, 0.96] (p < .001) | -0.50 [-0.66, -0.29] (p < .001) | -0.58 [-0.74, -0.37] (p < .001) | 0.49 [0.29, 0.65] (p < .001) | 0.55 [0.36, 0.69] (p < .001) | 0.10 [-0.13, 0.33] (p = 0.381) | -0.44 [-0.64, -0.19] (p < .001) |
| MPG | 0.93 [0.89, 0.96] (p < .001) | — | -0.48 [-0.65, -0.28] (p < .001) | -0.53 [-0.71, -0.30] (p < .001) | 0.53 [0.34, 0.68] (p < .001) | 0.55 [0.36, 0.69] (p < .001) | 0.05 [-0.19, 0.28] (p = 0.679) | -0.45 [-0.64, -0.20] (p < .001) |
| AVA_Echo_ | -0.50 [-0.66, -0.29] (p < .001) | -0.48 [-0.65, -0.28] (p < .001) | — | 0.33 [0.05, 0.56] (p = 0.018) | -0.39 [-0.58, -0.17] (p < .001) | -0.39 [-0.57, -0.17] (p < .001) | 0.02 [-0.22, 0.26] (p = 0.840) | 0.35 [0.07, 0.57] (p = 0.011) |
| AVA_CMR_ | -0.58 [-0.74, -0.37] (p < .001) | -0.53 [-0.71, -0.30] (p < .001) | 0.33 [0.05, 0.56] (p = 0.018) | — | -0.33 [-0.56, -0.06] (p = 0.014) | -0.48 [-0.67, -0.24] (p < .001) | 0.07 [-0.21, 0.33] (p = 0.640) | 0.44 [0.19, 0.64] (p < .001) |
| CaScore | 0.49 [0.29, 0.65] (p < .001) | 0.53 [0.34, 0.68] (p < .001) | -0.39 [-0.58, -0.17] (p < .001) | -0.33 [-0.56, -0.06] (p = 0.014) | — | 0.91 [0.86, 0.94] (p < .001) | 0.01 [-0.23, 0.24] (p = 0.955) | -0.62 [-0.76, -0.41] (p < .001) |
| ICV | 0.55 [0.36, 0.69] (p < .001) | 0.55 [0.36, 0.69] (p < .001) | -0.39 [-0.57, -0.17] (p < .001) | -0.48 [-0.67, -0.24] (p < .001) | 0.91 [0.86, 0.94] (p < .001) | — | -0.01 [-0.25, 0.22] (p = 0.904) | -0.53 [-0.70, -0.30] (p < .001) |
| IFV | 0.10 [-0.13, 0.33] (p = 0.381) | 0.05 [-0.19, 0.28] (p = 0.679) | 0.02 [-0.22, 0.26] (p = 0.840) | 0.07 [-0.21, 0.33] (p = 0.640) | 0.01 [-0.23, 0.24] (p = 0.955) | -0.01 [-0.25, 0.22] (p = 0.904) | — | -0.14 [-0.40, 0.13] (p = 0.297) |
| LMI | -0.44 [-0.64, -0.19] (p < .001) | -0.45 [-0.64, -0.20] (p < .001) | 0.35 [0.07, 0.57] (p = 0.011) | 0.44 [0.19, 0.64] (p < .001) | -0.62 [-0.76, -0.41] (p < .001) | -0.53 [-0.70, -0.30] (p < .001) | -0.14 [-0.40, 0.13] (p = 0.297) | — |

**Supplementary Table 4:**

|  | PCD-CT | EID-CT (Siemens) | EID-CT (Philips) |
| --- | --- | --- | --- |
| ECG-triggering phase | 60-90% | 60-90% | 40-90% |
| Tube voltage | 120 kVp | 100 kVp | 120 kVp |
| Tube current | automated | automated | automated |
| Slice thickness | 0.2 mm | 0.75 mm | 0.6 mm |
| Reconstruction increment | 0.1 mm | 0.4 mm | 0.3 mm |
| Reconstruction algorithm & strength | Quantum iterative reconstruction (4) | Quantum iterative reconstruction (4) | Iterative Model reconstruction (2) |
| Reconstruction Kernel | BV40 | BV36 | Cardiac routine |
